# Clinical Outcomes After Clopidogrel Versus Prasugrel Use in Endovascular Treatment of Unruptured Intracranial Aneurysms: A Propensity-Matched TriNetX Analysis

**DOI:** 10.64898/2026.10.04.26364712

**Authors:** Muhaison Hanifa Ibrahim, Simon Ammanuel, Tenzing Lhamo, Laura Stone McGuire

## Abstract

**Background:** Endovascular treatment of unruptured intracranial aneurysms (UIAs) often requires dual antiplatelet therapy with aspirin and a P2Y12 inhibitor. Clopidogrel remains widely used, but variable bioactivation and high on-treatment platelet reactivity have prompted interest in alternatives such as prasugrel. We compared thromboembolic and hemorrhagic outcomes after clopidogrel versus prasugrel use in endovascular UIA treatment.

**Methods:** We queried the TriNetX Global Collaborative Network for adults with UIA who underwent endovascular treatment between 2016 and 2025 and received aspirin plus clopidogrel or prasugrel. Cohorts were matched 1:1 using propensity score matching based on demographics and comorbidities. Outcomes were assessed at 6 months and 1 year and reported as odds ratios (ORs) with 95% confidence intervals (CIs).

**Results:** Among 15,496 adults, 711 matched pairs were included. At 6 months, no significant differences were observed for in-stent thrombosis (OR 0.77, 95% CI 0.38–1.57; p=0.47), cerebral infarction (OR 1.04, 95% CI 0.75–1.46; p=0.80), cerebral hemorrhage (OR 1.07, 95% CI 0.80–1.44; p=0.65), or gastrointestinal hemorrhage (OR 0.84, 95% CI 0.43–1.64; p=0.61). Findings remained nonsignificant at 1 year.

**Conclusions:** In this propensity-matched TriNetX analysis, clopidogrel and prasugrel were not associated with significant differences in thromboembolic or hemorrhagic outcomes after endovascular UIA treatment. Prasugrel may be a reasonable alternative P2Y12 inhibitor in selected patients, including those with suspected or documented clopidogrel non-response.

## INTRODUCTION

Unruptured intracranial aneurysms (UIAs) are common incidental neurovascular lesions, with prevalence estimates between 2 and 3% in the general population in the United States. Although the annual rupture risk for many UIAs is low, rupture typically results in aneurysmal subarachnoid hemorrhage, a catastrophic event associated with substantial mortality and long-term neurological disability.^1,2^ The decision to treat a UIA therefore requires individualized assessment of rupture risk - considering factors such as aneurysm size, location, morphology, and patient risk factors - balanced with overall life expectancy and procedural risk.^3^

Endovascular therapy has become a central treatment strategy for selected UIAs. Coil embolization, stent-assisted coiling, flow diversion, and intrasaccular devices have expanded the population of treatable aneurysms and shifted many centers away from open clipping for anatomically suitable lesions.^4,5^ However, the same endoluminal devices that improve aneurysm occlusion introduce a thrombogenic surface into the intracranial circulation. As a result, antiplatelet therapy is a critical adjunct for stent-assisted and flow-diversion procedures, where thromboembolic events and in-stent thrombosis can cause devastating ischemic injury.^6–8^

Aspirin plus clopidogrel has historically been the dominant DAPT regimen in neuroendovascular practice. Clopidogrel, however, is a prodrug that requires CYP2C19-mediated bioactivation, and loss-of-function alleles can reduce active metabolite formation and platelet inhibition.^9,10^ The Clinical Pharmacogenetics Implementation Consortium has strengthened genotype-guided recommendations for clopidogrel, including alternative antiplatelet strategies for patients with reduced CYP2C19 function in relevant vascular contexts.^9^ In neurointervention, high on-treatment platelet reactivity has been associated with thromboembolic complications, motivating many programs to use platelet function testing, genotype-guided selection, or empiric alternative P2Y12 inhibitors in selected patients.^11–16^

Prasugrel is a third-generation thienopyridine that provides more rapid and predictable P2Y12 inhibition than clopidogrel and is less dependent on CYP2C19 metabolism.^17^ In cardiology, prasugrel reduced ischemic events compared with clopidogrel in patients with acute coronary syndrome undergoing percutaneous coronary intervention, but at the cost of increased major bleeding in some subgroups, shaping its boxed warning and cautious use in patients with prior stroke or transient ischemic attack, older age, or low body weight.^18^ The relevance of this fixed-dose cardiology experience to neuroendovascular practice remains debated because contemporary neurointerventional protocols often use lower-dose prasugrel, PRU-guided dose adjustment, and carefully selected patient populations.^19^

Existing neuroendovascular studies suggest that prasugrel can reduce platelet reactivity and may lower thromboembolic complications without clearly increasing hemorrhage in selected aneurysm cohorts, but most reports are single-center, retrospective, or focused on specific devices and dosing strategies.^20–24^ Comparative data at scale remain limited, particularly for UIA patients treated in heterogeneous real-world settings. The TriNetX research network enables large, multi-institutional analyses of de-identified electronic health record data and provides a practical platform for evaluating comparative safety signals in lower-frequency outcomes.^25^ The objective of the present study was to compare thromboembolic and hemorrhagic outcomes after endovascular UIA treatment among patients receiving clopidogrel versus prasugrel as the P2Y12 component of DAPT. We used propensity score matching to balance measured baseline characteristics and evaluated clinically relevant outcomes at 6 months and 1 year after treatment.

## METHODS

### Study Design and Data Source

This retrospective cohort study used de-identified electronic health record data from the TriNetX Global Collaborative Network/Research Network, a federated research network comprising participating healthcare organizations. The study period extended between January 1, 2016, to December 31, 2025, and provided access to electronic medical records, including diagnoses, procedures, medications, and selected laboratory data, from approximately 130 million patients across 111 healthcare organizations. TriNetX data are de-identified before access by investigators. Analyses were conducted within the TriNetX analytics environment, and only aggregate results were exported for manuscript preparation.^26^

### Study Population

We identified adults (age >=18 years) with a diagnosis of UIA who underwent endovascular aneurysm treatment between January 1, 2016, and December 31, 2025. UIA was defined using ICD-10-CM code I67.1. Endovascular treatment was defined using CPT code 61624. The index date was defined as the date of the first qualifying endovascular treatment during the study period. Patients were excluded if they had evidence of a ruptured aneurysm or subarachnoid hemorrhage before or on the index date. Additional exclusions included missing demographic data or insufficient follow-up.

### Exposure Definition

The exposure of interest was the P2Y12 inhibitor used as part of DAPT. The clopidogrel cohort included patients with documented clopidogrel use within 6 months before or up to 6 months after the index endovascular procedure. The prasugrel cohort included patients with documented prasugrel use within the same peri-procedural window. Medication records in TriNetX reflect orders, prescriptions, or administrations and do not guarantee outpatient adherence. The cohort was defined by endovascular aneurysm treatment and antiplatelet exposure rather than by separate device-specific intracranial stent or flow-diverter procedural codes.

### Covariates and Propensity Score Matching

To reduce confounding by indication and measured baseline differences, clopidogrel and prasugrel cohorts were matched 1:1 using propensity score matching within TriNetX. The propensity model included age, sex, race, ethnicity, body mass index, and clinically relevant comorbidities, including hypertension, diabetes mellitus, hyperlipidemia, atrial fibrillation, chronic kidney disease, coronary artery disease, peripheral artery disease, and tobacco use. Covariate balance was assessed using standardized mean differences (SMDs), with an absolute SMD <=0.10 considered acceptable balance.

### Outcomes

Outcomes were assessed at 6 months and 1 year after the index procedure. The prespecified outcomes were in-stent thrombosis, cerebral infarction, cerebral hemorrhage, and gastrointestinal hemorrhage. Cerebral infarction was identified using ICD-10-CM code I63. Cerebral hemorrhage was identified using ICD-10-CM codes I60 and I61 when recorded after the index date. Gastrointestinal hemorrhage was identified using ICD-10-CM codes K92.0, K92.1, K92.2, and K62.5. In-stent thrombosis was operationalized as a coded device-related thrombotic or stenotic complication occurring after endovascular aneurysm treatment. Specifically, this outcome was ascertained using ICD-10-CM T82 family codes capturing complications, thrombosis, or stenosis of cardiac and vascular prosthetic devices, implants, and grafts, including T82.85, T82.858, T82.86, T82.868, and related descendant or encounter-specific codes available within TriNetX. To increase clinical relevance to the treated UIA population, these codes were evaluated only among patients meeting the endovascular UIA treatment cohort definition and only when newly recorded after the index procedure within the specified follow-up window.

### Statistical Analysis

Baseline characteristics were summarized before and after matching using means with standard deviations or medians with interquartile ranges for continuous variables and counts with percentages for categorical variables, as available from TriNetX. Between-cohort outcome associations were reported as odds ratios (ORs) with 95% confidence intervals (CIs); ORs are presented for clopidogrel relative to prasugrel unless otherwise specified. Statistical significance was defined as a two-sided p<0.05. No correction for multiple comparisons was applied, and findings should be interpreted as exploratory and hypothesis-generating.

## RESULTS

### Cohort Selection and Baseline Characteristics

A total of 15,496 adult patients met the initial study criteria for endovascular treatment of UIAs during the study period. After applying antiplatelet exposure criteria, 14,785 patients were assigned to the clopidogrel cohort and 711 patients to the prasugrel cohort before propensity score matching. Following 1:1 propensity score matching, 711 patients remained in each cohort for comparative analysis.

Before matching, several baseline differences were observed between the clopidogrel and prasugrel cohorts. Patients in the clopidogrel cohort were slightly younger at the index encounter than those in the prasugrel cohort (58.8 ± 13.3 vs. 60.6 ± 12.7 years, p=0.0005), although current age was similar between groups (65.6 ± 13.6 vs. 65.4 ± 13.0 years, p=0.8171). The clopidogrel cohort also had a lower proportion of female patients (75.7% vs. 81.3%, p=0.0006). Additional pre-match differences were observed in selected racial and ethnic categories, as well as in hypertension, hyperlipidemia, atherosclerotic cardiovascular disease, and chronic kidney disease.

Propensity score matching substantially improved balance across measured demographic and clinical covariates. In the matched cohorts, current age was similar between clopidogrel and prasugrel patients (65.1 ± 13.4 vs. 65.4 ± 13.0 years, p=0.6228), as was age at index encounter (60.5 ± 13.3 vs. 60.6 ± 12.7 years, p=0.8402). Female patients comprised 80.7% of the clopidogrel cohort and 81.3% of the prasugrel cohort after matching (p=0.7868). Baseline racial and ethnic distributions were also comparable between groups after matching.

Comorbidities were similarly balanced after matching. There were no statistically significant differences between the matched clopidogrel and prasugrel cohorts in hypertension (65.5% vs. 65.5%, p=1.0000), hyperlipidemia (38.3% vs. 39.7%, p=0.5866), nicotine dependence or tobacco use (28.4% vs. 27.7%, p=0.7679), atrial fibrillation (7.5% vs. 8.4%, p=0.4925), chronic kidney disease (7.2% vs. 8.9%, p=0.2413), other peripheral vascular disease (6.8% vs. 7.2%, p=0.7546), or pulmonary embolism (1.5% vs. 1.7%, p=0.8335). Atherosclerotic cardiovascular disease remained numerically more frequent in the prasugrel cohort after matching, but this difference did not reach statistical significance (11.7% vs. 14.9%, p=0.0724). Overall, propensity score matching generated well-balanced treatment cohorts for subsequent outcome comparisons.

### Six-Month Outcomes

At 6 months after endovascular UIA treatment, no statistically significant differences were observed between clopidogrel and prasugrel in any prespecified thromboembolic or hemorrhagic outcome (Table 2). In-stent thrombosis did not differ significantly between groups (OR 0.77, 95% CI 0.38-1.57; p=0.47). Cerebral infarction (OR 1.04, 95% CI 0.75-1.46; p=0.80), cerebral hemorrhage (OR 1.07, 95% CI 0.80-1.44; p=0.65), and gastrointestinal hemorrhage (OR 0.84, 95% CI 0.43-1.64; p=0.61) were also not significantly different.

**Table 1.** Baseline characteristics after propensity score matching.

| Characteristic | Clopidogrel (n=711) | Prasugrel (n=711) | P-value | \|SMD\| |
| --- | --- | --- | --- | --- |
| **Demographics** |  |  |  |  |
| **Current age, years** | 65.1 ± 13.4; median 66<br>(IQR 17) | 65.4 ± 13.0; median 67<br>(IQR 17) | 0.6228 | 0.023 |
| **Age at index, years** | 60.5 ± 13.3; median 62<br>(IQR 18) | 60.6 ± 12.7; median 62<br>(IQR 18) | 0.8402 | 0.008 |
| **Sex** |  |  |  |  |
| **Female** | 574 (80.7%) | 578 (81.3%) | 0.7868 | 0.014 |
| **Male** | 137 (19.3%) | 133 (18.7%) | 0.7868 | 0.014 |
| **Unknown gender** | 0 (0.0%) | 0 (0.0%) | — | 0.000 |
| **Ethnicity** |  |  |  |  |
| **Not Hispanic or Latino** | 531 (74.7%) | 527 (74.1%) | 0.8080 | 0.013 |
| **Hispanic or Latino** | 37 (5.2%) | 36 (5.1%) | 0.9044 | 0.006 |
| **Unknown ethnicity** | 143 (20.1%) | 148 (20.8%) | 0.7424 | 0.017 |
| **Race** |  |  |  |  |
| **White** | 516 (72.6%) | 507 (71.3%) | 0.5953 | 0.028 |
| **Black or African American** | 95 (13.4%) | 114 (16.0%) | 0.1547 | 0.076 |
| **Asian** | 34 (4.8%) | 25 (3.5%) | 0.2314 | 0.064 |
| **American Indian or Alaska Native** | 10 (1.4%) | 10 (1.4%) | 1.0000 | 0.000 |
| **Native Hawaiian or**<br>**Other Pacific**<br>**Islander** | 0 (0.0%) | 0 (0.0%) | — | 0.000 |
| **Other race** | 10 (1.4%) | 13 (1.8%) | 0.5283 | 0.033 |
| **Unknown race** | 54 (7.6%) | 47 (6.6%) | 0.4679 | 0.038 |
| **Comorbidities** |  |  |  |  |
| **Hypertension** | 466 (65.5%) | 466 (65.5%) | 1.0000 | 0.000 |
| **Hyperlipidemia** | 272 (38.3%) | 282 (39.7%) | 0.5866 | 0.029 |
| **Nicotine dependence** | 202 (28.4%) | 197 (27.7%) | 0.7679 | 0.016 |
| **Atherosclerotic heart**<br>**disease of native**<br>**coronary artery** | 83 (11.7%) | 106 (14.9%) | 0.0724 | 0.095 |
| **Atrial**<br>**fibrillation/flutter** | 53 (7.5%) | 60 (8.4%) | 0.4925 | 0.036 |
| **Chronic kidney**<br>**disease** | 51 (7.2%) | 63 (8.9%) | 0.2413 | 0.062 |
| **Other peripheral**<br>**vascular disease** | 48 (6.8%) | 51 (7.2%) | 0.7546 | 0.017 |
| **Pulmonary embolism** | 11 (1.5%) | 12 (1.7%) | 0.8335 | 0.011 |

*Values are mean ± SD; median (IQR), or n (%). SMD indicates standardized mean difference.*

**Table 2.**
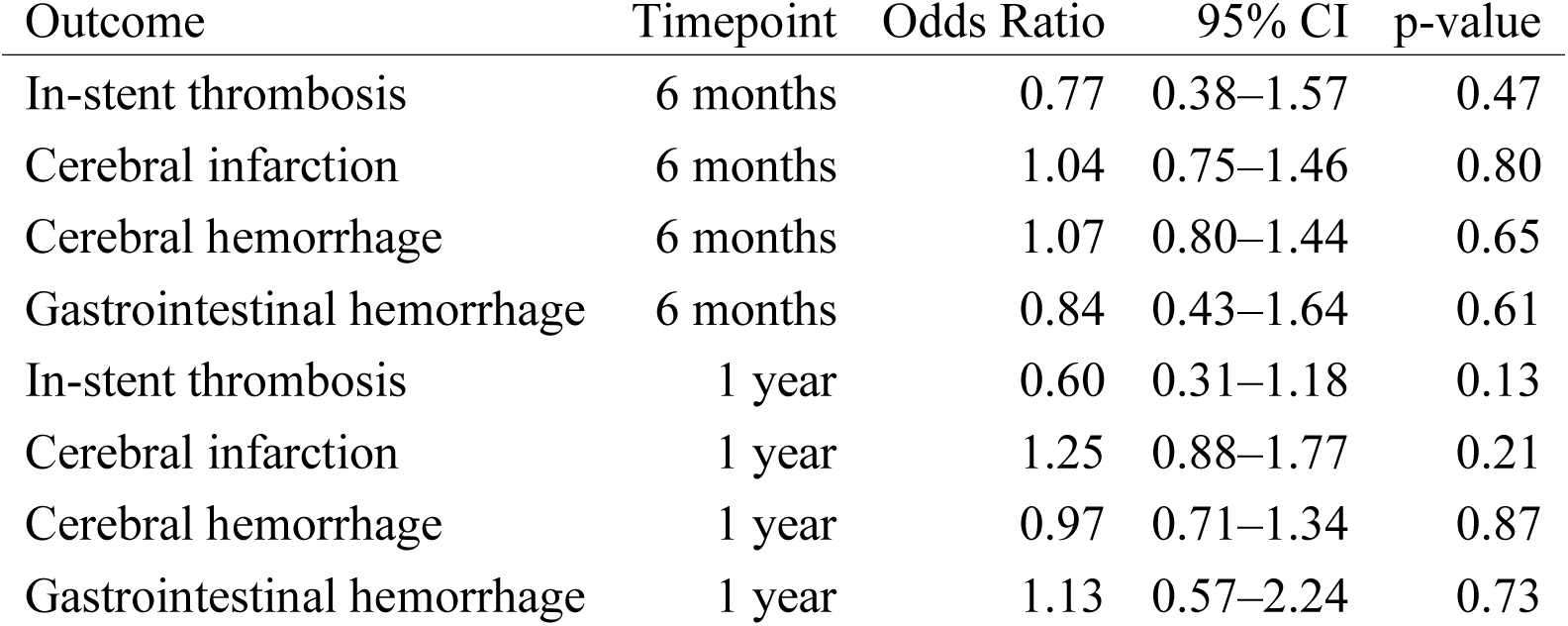
Outcomes after propensity score matching (odds ratios for clopidogrel relative to prasugrel)

### One-Year Outcomes

Findings remained nonsignificant at 1 year (Table 2). There were no statistically significant differences in in-stent thrombosis (OR 0.60, 95% CI 0.31-1.18; p=0.13), cerebral infarction (OR 1.25, 95% CI 0.88-1.77; p=0.21), cerebral hemorrhage (OR 0.97, 95% CI 0.71-1.34; p=0.87), or gastrointestinal hemorrhage (OR 1.13, 95% CI 0.57-2.24; p=0.73).

In-stent thrombosis was identified using ICD-10-CM T82-family codes for device-related thrombotic or stenotic complications after the index endovascular aneurysm procedure and was not angiographically adjudicated.

## DISCUSSION

In this propensity score-matched TriNetX analysis of adults undergoing endovascular treatment for UIAs, clopidogrel and prasugrel were not associated with statistically significant differences in in-stent thrombosis, cerebral infarction, cerebral hemorrhage, or gastrointestinal hemorrhage at 6 months or 1 year. After matching, 711 patients in each treatment cohort were included, making this a large real-world comparison of P2Y12 inhibitor selection in the UIA endovascular population. The findings support the clinical plausibility of prasugrel as an alternative to clopidogrel in selected patients, while also emphasizing that the present design cannot establish equivalence, noninferiority, or superiority.

The absence of a significant difference in thromboembolic outcomes should be interpreted in the context of clopidogrel pharmacogenomics and real-world prescribing. Clopidogrel non-response is clinically important but not universal. In a population-level EHR cohort, many clopidogrel-treated patients likely had adequate platelet inhibition, whereas prasugrel may have been preferentially used in patients perceived to be at higher thrombotic risk or in those with known or suspected clopidogrel resistance. These competing factors may attenuate measurable differences between agents, particularly when platelet reactivity, CYP2C19 genotype, aneurysm morphology, device type, and operator-level practice are unavailable.

The hemorrhagic findings are clinically relevant because prasugrel has historically carried concern for increased bleeding based largely on cardiology trial experience. In the present analysis, cerebral hemorrhage and gastrointestinal hemorrhage were not significantly different between cohorts at either timepoint. This aligns with a growing neurointerventional literature suggesting that lower-dose or platelet function-guided prasugrel strategies may provide potent

P2Y12 inhibition without a clear excess of hemorrhagic complications in carefully selected patients.^14,15^ Nevertheless, the confidence intervals for several outcomes remain wide, and rare complications, such as in-stent thrombosis or intracranial hemorrhage, require cautious interpretation.

These results have practical implications for antiplatelet selection. For patients with adequate clopidogrel response, clopidogrel remains a familiar, widely used, and broadly available agent. For patients with high on-treatment platelet reactivity, CYP2C19 loss-of-function alleles, prior clopidogrel failure, or institutional protocols favoring alternative P2Y12 inhibition, prasugrel appears to be a defensible option based on the present matched analysis. Accordingly, the findings should not be framed as demonstrating therapeutic equivalence; rather, they suggest no detectable safety signal against prasugrel in the measured outcomes within the limitations of the database.

The study also highlights the need for more granular comparative research. Future studies should incorporate procedure type, device platform, aneurysm size and location, antiplatelet loading and maintenance doses, DAPT duration, adherence, platelet function testing, CYP2C19 genotype, and operator or center-level protocols. Prospective pragmatic studies or registry-based randomized trials would be particularly useful for determining whether prasugrel offers a clinically meaningful advantage in biologically defined subgroups, such as clopidogrel intermediate or poor metabolizers.

### Limitations

This study has several limitations. First, TriNetX analyses depend on diagnosis, procedure, and medication coding, which can lead to misclassification of UIA status, procedure type, medication exposure, and outcomes. Second, claims and EHR data do not provide complete procedural granularity, including aneurysm size, location, morphology, rupture-risk profile, device type, number of devices, technical success, intraprocedural complications, or operator experience. Third, the medication exposure definition captures documentation of prescriptions, orders, or administrations, but it cannot confirm outpatient adherence, exact loading dose, maintenance dose, dose adaptation to platelet reactivity testing, DAPT duration, switching between agents, or perioperative interruptions.

Additionally, in-stent thrombosis was identified using ICD-10-CM T82 family codes for device-related thrombotic or stenotic complications rather than angiographic adjudication or intracranial stent-specific procedural review, which may introduce outcome misclassification.

Fourth, although propensity score matching balances measured baseline covariates, residual confounding by unmeasured factors remains likely. In particular, prasugrel may be preferentially prescribed to patients with clopidogrel non-response or perceived higher thrombotic risk, while clinicians may avoid prasugrel in patients perceived to be at higher bleeding risk. Fifth, the analysis does not include platelet function testing or CYP2C19 genotype data, both of which are central to the biological rationale for choosing prasugrel over clopidogrel. Sixth, outcome timing is based on follow-up windows rather than adjudicated time-to-event data; furthermore, recurrent events, event severity, functional outcomes, retreatment, aneurysm occlusion, and mortality were not assessed.

Finally, nonsignificant p-values should not be interpreted as equivalence. The study was not designed with a prespecified equivalence or noninferiority margin, and several confidence intervals remain compatible with clinically meaningful differences in either direction. These findings should therefore be considered hypothesis-generating and should be validated in prospective cohorts with standardized antiplatelet protocols and adjudicated outcomes.

## CONCLUSIONS

In this propensity score-matched TriNetX analysis of adults undergoing endovascular treatment for UIAs, clopidogrel and prasugrel were not associated with statistically significant differences in in-stent thrombosis, cerebral infarction, cerebral hemorrhage, or gastrointestinal hemorrhage at 6 months or 1 year. The findings support prasugrel as a reasonable alternative P2Y12 inhibitor in selected patients, particularly when clopidogrel non-response is suspected or documented.

Prospective studies incorporating antiplatelet dose, DAPT duration, platelet function testing, CYP2C19 genotype, aneurysm anatomy, and device-level procedural data are needed to define optimal P2Y12 selection in neuroendovascular UIA treatment.

## Data Availability

Available upon request

## Ethical approval

Not applicable; this study used de-identified electronic health record data from the TriNetX Global Collaborative Network.

## Consent to participate

Individual informed consent was not obtained because investigators did not access identifiable patient-level information and only aggregate results were exported.

## Acknowledgement

None

## REFERENCES

1. Rinkel GJ, Djibuti M, Algra A, et al. Prevalence and risk of rupture of intracranial aneurysms: a systematic review. Stroke 1998; 29: 251–256.

2. van Gijn J, Kerr RS, Rinkel GJE. Subarachnoid haemorrhage. Lancet 2007; 369: 306–318.

3. Algra AM, Greving JP, de Winkel J, et al. Development of the SAFETEA Scores for Predicting Risks of Complications of Preventive Endovascular or Microneurosurgical Intracranial Aneurysm Occlusion. Neurology 2022; 99: e1725–e1737.

4. Brinjikji W, Rabinstein AA, Nasr DM, et al. Better Outcomes with Treatment by Coiling Relative to Clipping of Unruptured Intracranial Aneurysms in the United States, 2001–2008. AJNR Am J Neuroradiol 2011; 32: 1071–1075.

5. Pierot L, Wakhloo AK. Endovascular treatment of intracranial aneurysms: current status. Stroke 2013; 44: 2046–2054.

6. Biondi A, Janardhan V, Katz JM, et al. Neuroform stent-assisted coil embolization of wide-neck intracranial aneurysms: strategies in stent deployment and midterm follow-up. Neurosurgery 2007; 61: 460–468; discussion 468-469.

7. Brinjikji W, Lanzino G, Cloft HJ, et al. Risk Factors for Ischemic Complications following Pipeline Embolization Device Treatment of Intracranial Aneurysms: Results from the IntrePED Study. American Journal of Neuroradiology 2016; 37: 1673–1678.

8. Schirmer CM, Bulsara KR, Al-Mufti F, et al. Antiplatelets and antithrombotics in neurointerventional procedures: Guideline update. J Neurointerv Surg 2023; 15: 1155–1162.

9. Lee CR, Luzum JA, Sangkuhl K, et al. Clinical Pharmacogenetics Implementation Consortium Guideline for CYP2C19 Genotype and Clopidogrel Therapy: 2022 Update. Clin Pharmacol Ther 2022; 112: 959–967.

10. Mega JL, Close SL, Wiviott SD, et al. Cytochrome p-450 polymorphisms and response to clopidogrel. N Engl J Med 2009; 360: 354–362.

11. Desta Z, Zhao X, Shin J-G, et al. Clinical significance of the cytochrome P450 2C19 genetic polymorphism. Clin Pharmacokinet 2002; 41: 913–958.

12. Hwang G, Huh W, Lee JS, et al. Standard vs Modified Antiplatelet Preparation for Preventing Thromboembolic Events in Patients With High On-Treatment Platelet Reactivity Undergoing Coil Embolization for an Unruptured Intracranial Aneurysm: A Randomized Clinical Trial. JAMA Neurol 2015; 72: 764–772.

13. Delgado Almandoz JE, Crandall BM, Scholz JM, et al. Last-recorded P2Y12 reaction units value is strongly associated with thromboembolic and hemorrhagic complications occurring up to 6 months after treatment in patients with cerebral aneurysms treated with the pipeline embolization device. AJNR Am J Neuroradiol 2014; 35: 128–135.

14. Mitchell DL, McGuire LS, Khalid SI, et al. Assessing the efficacy of VerifyNow platelet-function testing in predicting postoperative thromboembolic complications of neuroendovascular surgery: A systematic review and meta-analysis (part 1). Interv Neuroradiol 2026; 32: 908–920.

15. Mitchell DL, McGuire LS, Khalid SI, et al. Assessing the efficacy of VerifyNow platelet-function testing in predicting postoperative hemorrhagic complications of neuroendovascular surgery: A systematic review and meta-analysis (part 2). Interv Neuroradiol 2026; 32: 921–930.

16. Fifi JT, Brockington C, Narang J, et al. Clopidogrel resistance is associated with thromboembolic complications in patients undergoing neurovascular stenting. AJNR Am J Neuroradiol 2013; 34: 716–720.

17. Mega JL, Close SL, Wiviott SD, et al. Cytochrome P450 genetic polymorphisms and the response to prasugrel: relationship to pharmacokinetic, pharmacodynamic, and clinical outcomes. Circulation 2009; 119: 2553–2560.

18. Wiviott SD, Braunwald E, McCabe CH, et al. Prasugrel versus clopidogrel in patients with acute coronary syndromes. N Engl J Med 2007; 357: 2001–2015.

19. Ibrahim MH, Lee B, McGrath J, et al. Reassessing prasugrel’s boxed warning: a review of dose adjustment, platelet function testing, and platelet reactivity unit (PRU)-guided strategies in neurointervention. J Neurointerv Surg 2026; jnis-2026-025144.

20. Flynn LM, Mohamed E, Dobbs N, et al. Safety of dual antiplatelet therapy using aspirin and low-dose Prasugrel with platelet reactivity testing in flow diverter treatment of intracranial aneurysms. Interv Neuroradiol 2026; 32: 460–465.

21. Sedat J, Chau Y, Gaudart J, et al. Prasugrel versus clopidogrel in stent-assisted coil embolization of unruptured intracranial aneurysms. Interv Neuroradiol 2017; 23: 52–59.

22. Ha EJ, Cho WS, Kim JE, et al. Prophylactic Antiplatelet Medication in Endovascular Treatment of Intracranial Aneurysms: Low-Dose Prasugrel versus Clopidogrel. AJNR Am J Neuroradiol 2016; 37: 2060–2065.

23. Cagnazzo F, Perrini P, Lefevre P-H, et al. Comparison of Prasugrel and Clopidogrel Used as Antiplatelet Medication for Endovascular Treatment of Unruptured Intracranial Aneurysms: A Meta-Analysis. AJNR Am J Neuroradiol 2019; 40: 681–686.

24. Hohenstatt S, Saatci I, Jesser J, et al. Prasugrel Single Antiplatelet Therapy versus Aspirin and Clopidogrel Dual Antiplatelet Therapy for Flow Diverter Treatment for Cerebral Aneurysms: A Retrospective Multicenter Study. AJNR Am J Neuroradiol 2024; 45: 592–598.

25. Palchuk MB, London JW, Perez-Rey D, et al. A global federated real-world data and analytics platform for research. JAMIA Open 2023; 6: ooad035.

26. TriNetX. Publication Guidelines. TriNetX, https://trinetx.com/publication/trinetx-publication-guidelines/ (accessed 29 July 2026).

